# Convalescent Plasma Use in the United States was inversely correlated with COVID-19 Mortality: Did Plasma Hesitancy cost lives?

**DOI:** 10.1101/2021.04.07.21255089

**Authors:** Arturo Casadevall, Quigly Dragotakes, Patrick W. Johnson, Jonathon W. Senefeld, Steven A. Klassen, R. Scott Wright, Michael J Joyner, Nigel Paneth, Rickey E. Carter

## Abstract

**Background:** The US Food and Drug Administration authorized Convalescent Plasma (CCP) therapy for hospitalized COVID-19 patients via the Expanded Access Program (EAP) and the Emergency Use Authorization (EUA), leading to use in about 500,000 patients during the first year of the pandemic for the US.

**Methods:** We tracked the number of CCP units dispensed to hospitals by blood banking organizations and correlated that usage with hospital admission and mortality data.

**Results:** CCP usage per admission peaked in Fall 2020, with more than 40% of inpatients estimated to have received CCP between late September and early November 2020. However, after randomized controlled trials failed to show a reduction in mortality, CCP usage per admission declined steadily to a nadir of less than 10% in March 2021. We found a strong inverse correlation (r = −0.52, P = 0.002) between CCP usage per hospital admission and deaths occurring two weeks after admission, and this finding was robust to examination of deaths taking place one, two or three weeks after admission. Changes in the number of hospital admissions, SARS-CoV-2 variants, and age of patients could not explain these findings. The retreat from CCP usage might have resulted in as many as 29,000 excess deaths from mid-November 2020 to February 2021.

**Conclusions:** A strong inverse correlation between CCP use and mortality per admission in the USA provides population level evidence consistent with the notion that CCP reduces mortality in COVID-19 and suggests that the recent decline in usage could have resulted in excess deaths.

## Introduction

In the Spring of 2020, the United States embarked on a historic and unprecedented deployment of plasma derived from patients who survived COVID-19 (COVID-19 Convalescent Plasma (CCP)) for treatment of the disease, and one-year into this effort more than 500,000 individuals have been treated. The synergism created by the lack of effective alternate therapies, a plentiful supply of plasma from an efficient and high-capacity blood banking network, motivated donors and strong community partners fueled this deployment. Sensible US FDA regulatory oversight was provided first by its Expanded Access Program (EAP) in partnership with the Mayo Clinic, with first transfusion on early April 2020,^1^ and then by its Emergency Use Authorization (EUA) of August 23, 2020, both of which restricted CCP use to hospitalized patients ^2^.

The demonstration by the summer of 2020 that CCP was safe ^3,4^, that antibody in plasma correlated with survival in people treated before ventilation ^5^ along with initial suggestions of efficacy ^6-8^, fueled interest in and use of this product. However, the interpretation of the potential efficacy of CCP is complex as many of the positive findings arose through post hoc examinations and subgroup comparisons. Nonetheless, the use of CCP rose rapidly without the ideal evidence base of efficacy from randomized controlled clinical trials (RCT), since early RCTs though generally trending positively, were unsatisfactory, mostly due to small size or premature termination as the epidemic abated in the early surge regions ^9^. Later in the pandemic several larger RCTs reported no mortality benefit ^10-12^, raising doubts as to CCP efficacy. However, these latter trials were undertaken in hospitalized patients treated late in the course of disease and some used plasma with variable antibody levels ^9^, and contrasted with a highly successful trial in elderly patients treated within 3 days of illness onset prior to hospitalization ^13^. Despite potential explanations for the negative studies, the results of these studies were sometimes accompanied by editorials that reinforced message of futility with the British Medical Journal calling CCP ‘ineffective’ ^14^, Nature Biotechnology reported that CCP fell ‘flat’ ^15^ and JAMA published a meta-analysis of RCT concluding that there was no evidence of benefit from CCP therapy ^16^. On February 17, 2021 the *Wall Street Journal* reported that Mount Sinai Hospital, which had been a leader in deploying CCP and reported early encouraging results ^8^, had stopped using plasma in patients with COVID-19, and the report specifically mentioned the negative results from CCP RCTs in this decision ^17^.

On March 13, 2021 the New York Times reported that COVID-19 mortality remained high with nearly 1,500 daily deaths despite a drop in the number of new infections since earlier in the year ^18^. This finding was surprising in light of an apparent reduction in the mortality of hospitalized patients as the epidemic progressed, thought to be from improved management of the disease as clinical experience grew ^19^. Analyzing weekly reports from the blood banking industry, we noted that plasma use was on the decline, based on the ratio of units dispensed to hospital admissions. The increase in mortality combined with the reduction in CCP use led us to hypothesize first, that the two phenomena were related, and second, that the decline in CCP use reflected reduced use following the disappointing trial findings. The blood banking network maintains careful and complete records for every blood product unit used including time, date and geographic provenance and destination, providing a virtually complete record of trends in CCP use for the US. We therefore examined the use of CCP units as a function of time, assessing the relationship of CCP use to COVID-19 mortality, denominating both plasma units and deaths to hospital admissions. The aim of the study was to determine whether the reduction in plasma use was associated with any change in the pattern of mortality seen in patients hospitalized for COVID-19.

## Materials and Methods

### Convalescent Plasma Usage

CCP usage was inferred from the distribution of plasma units to hospitals in the USA from data obtained from Blood Centers of America (BCA, West Warwick, RI). Data fields included collections, distributions to hospitals, distributions to research or other use. This file consolidated all the reports from regional blood bank reports and provided a total of collected units and units distributed to hospitals. The data file did not have information on whether a unit was actually transfused but BCA can infer usage from hospital re-ordering information and there has been a strong correlation between the total number of units shipped to a hospital and the units transfused by that hospital. Hence, the CCP units dispensed to hospitals represent a reasonable proxy value for the total number of units being transfused to patients. To validate this assumption, we compared the numbers of plasma units dispensed to those used by the EAP. There was a powerful and significant correlation between the weekly counts of units distributed in the United States and those used to treat patients as part of the EAP between April 06 and August 23, 2020. (Spearman rho = 0.953, P<0.001) (Figure S1). Units transfused in the EAP were reported by providers as part of the official case report forms and each transfusion could comprise one or two units.

### Admission and mortality data

For population level data on COVID-19 admissions and mortality we relied on publicly available databases. Specifically, we used information from the Our World in Data (OWID) (https://ourworldindata.org/coronavirus) database. Data used for this analysis were downloaded on March 18, 2021 and are available as Supplemental Table S1. We confirmed these findings using Centers for Disease Control (CDC, Atlanta, GA) data on admissions and deaths https://covid.cdc.gov/covid-data-tracker/#new-hospital-admissions and https://covid.cdc.gov/covid-data-tracker/#trends_dailytrendsdeaths. CDC data were downloaded on 3/31/2021, and are available as Supplemental Table S2.

### Statistical analysis

Preliminary descriptive analyses were used to explore the associations of the ratio of number of CCP units dispersed to the number of hospitalizations (CCP utilization ratio) with the ratio of national deaths to national admissions, the latter being a reasonable proxy for the case fatality rate. No individual-level data were available to link the mortality events directly to the individuals hospitalized to permit a calculation of the true case-fatality rate. To address this limitation, the mortality counts reported by the CDC were shifted to better align the deaths with the admitted patients. Since the overwhelming majority of COVID-19 deaths occur in hospitals ^20,21^, since CCP is only authorized for use in hospitals, and since death generally occurs a few days to weeks after admission, mortality was adjusted for the time lag between admission and death. The median time between admission and death has been reported as 9 days in the US ^22^ and 6.7 days in Belgium ^23^. For the analysis, which was based on weekly aggregated data, a two-week shift was selected to align the mortality with the median and upper quartile estimates in these reports. The Pearson correlation coefficient was used to describe the relationship of the CCP utilization ratio with CFR. To further define this relationship, a linear statistical model was used to regress the utilization ratio onto CFR. This statistical model was weighted by the number of hospitalizations per week. The fit of the model was examined using standard residual-based diagnostic plots and the fit was deemed acceptable using only a linear fit of the CCP utilization ratio.

Three in silico scenarios were created to summarize the effect of alterations to the CCP utilization ratio using the fitted model. In scenario 1, the effect of maintenance of plasma usage was considered. To define maintenance of use, a weighted average of the utilization ratio over the months of August through September 2020 was estimated. This value was then used to estimate the number of deaths that would be expected to occur throughout the study period (admissions starting 8/3/2020 – 2/22/2021). In scenario 2 a constant 50% CCP utilization ratio was set over the entire study period. The utilization rate was approximately the value observed in the early October 2020 period. A final scenario estimated the CFR that may have been observed had CCP not been used at all (i.e., the y-intercept from the model). Model contrasts were used to estimate the change in expected deaths among these scenarios in addition to a fourth condition – the actual number of events reported by the CDC. Values are summarized based on the observed number of hospitalized patients over the study period along with the same values indexed into expected mortality events per 1000 hospitalizations. Pointwise confidence bands for each scenario were obtained by multiplying the model-predicted CFR, and its associated 95% confidence interval, by the number of hospitalizations per week. Cumulative summations were used to describe the differences in expected deaths over the entire study period. This analysis was repeated independently a third time using a weighted average of utilization ratio of the months October-November on a separate database (OWID) to investigate stability between reporting bodies. As a final method for estimating the overall effect of the changes in CCP utilization, the CDC data were grouped into two time periods representing relative use. The difference in the CFR was used to estimate the expected value for the changes in the expected number of deaths.

Statistical analyses were conducted using R version 3.6.2.^24^ 95% confidence intervals and two-sided p-values were used to summarize association and test for significance at the alpha=0.05 level of significance, respectively.

## Results

### Convalescent plasma use

The FDA first allowed compassionate use of CCP on a case-by-case basis in late March 2020, but very quickly initiated the Expanded Access Program in early April 2020. The EAP was, in effect, a registry of all CCP use in the US from April to August 2020 and led to a sharp rise in CCP use. The findings of the EAP, which established that CCP was as relatively safe ^3,4^, and that high antibody titer was associated with lower mortality in unventilated plasma recipients ^5^, were major considerations behind the Emergency Use Authorization of August 23, 2020, which broadened its use. Distribution of CCP to hospitals rose to 25,000 - 30,000 weekly units by the December 2020 to January 2021 time period, but this rise in plasma distribution largely reflected the great increase in hospital admissions for COVID in those months. (Figure 1). When CCP distributions are analyzed as a function of the number of new hospital admissions per week, peak utilization per capita occurred much earlier, in early October 2020 and declined sharply in the following months (Figure 2).

**Figure 1.**
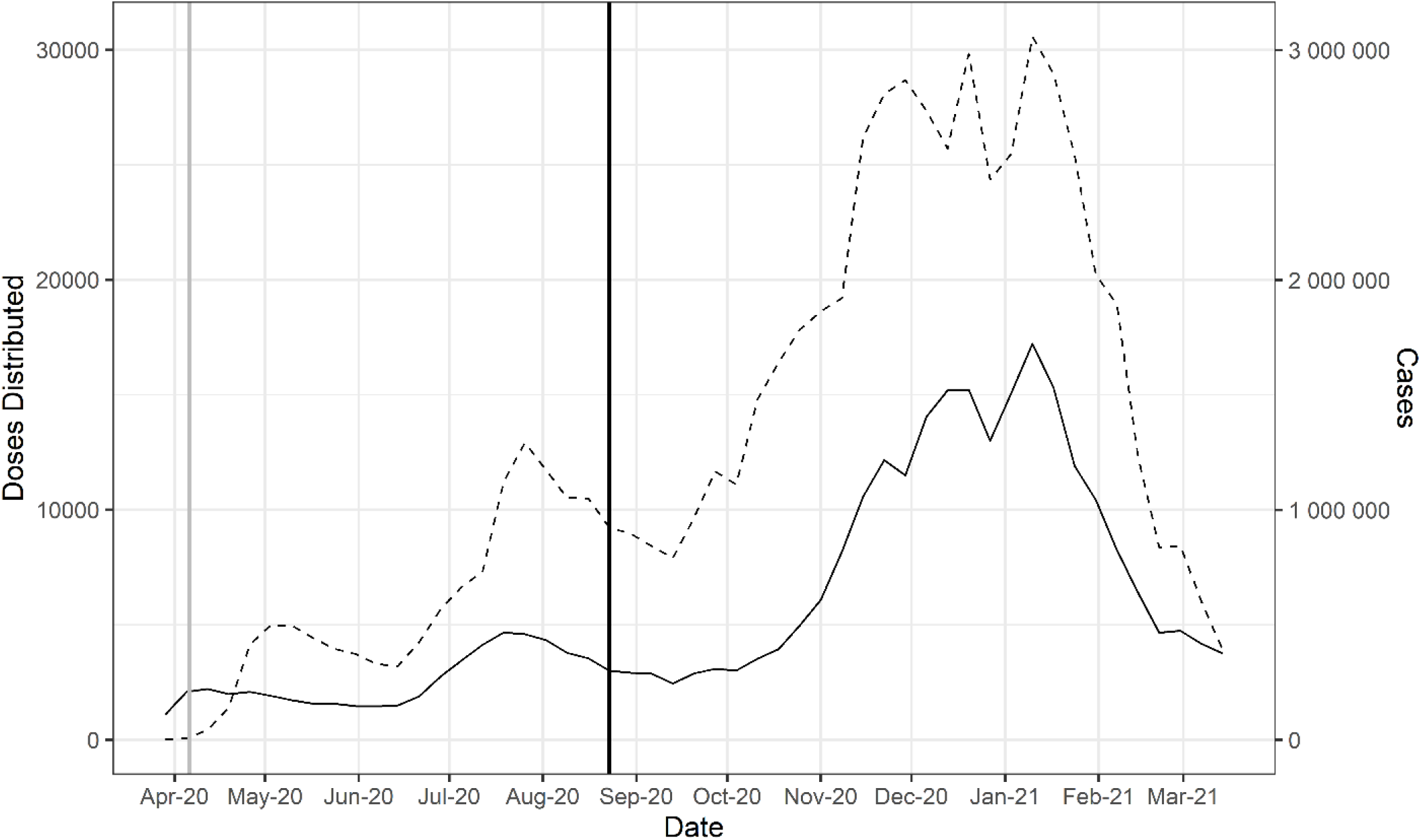
Doses of CCP distributed in the United States by the American Red Cross and American Blood Centers (dashed) and total COVID-19 cases in the United States reported in OWID (solid). The vertical black line marks August 23, 2020 when the FDA announced that Emergency Use Authorization for CCP in the USA. The vertical gray line marks April 4, 2020 as the start of the Emergency Access Program.

**Figure 2.**
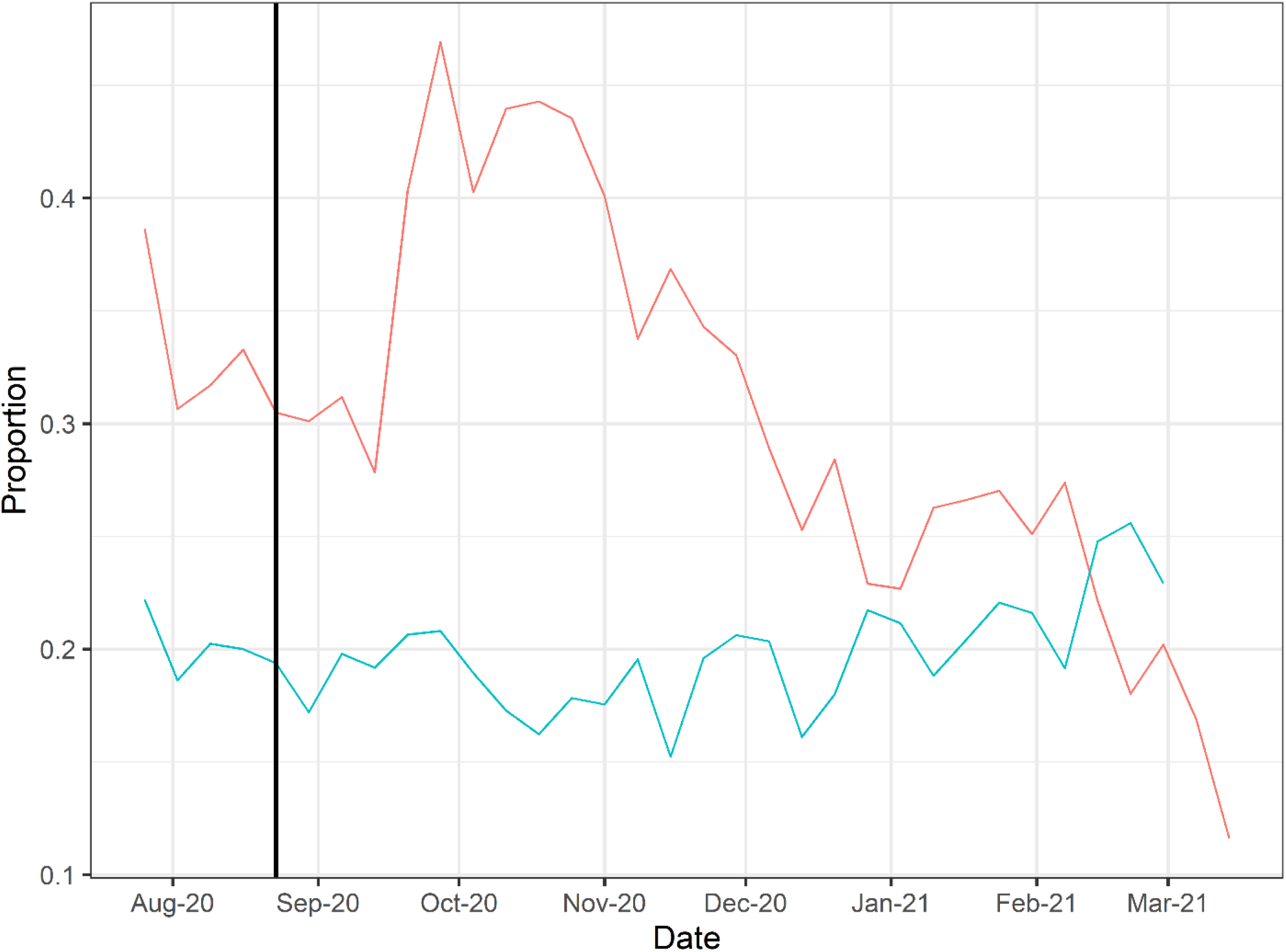
Doses of CCP per hospital admission (red) and mortality calculated as deaths per hospital admission (green). To account for time between admission to death, deaths from two weeks after admission are used to calculate mortality. The vertical line marks August 23, 2020 when the FDA announced that Emergency Use Authorization for CCP in the USA.

### Correlation between CCP and mortality

To explore whether there was a relationship between CCP distribution in the USA and mortality we first compared the doses per patient versus reported COVID-19 deaths per hospital admission from publicly available databases (Figure 2). The comparison of curves showed a trough in deaths per admission coinciding with the peak of CCP usage per admission. A plot of mortality versus doses per hospitalized patient using mortality per admission data from the OWID database revealed a strong negative correlation (Pearson’s correlation coefficient of −0.52 with P = 0.002) (Figure 3). To account for lags in the reporting of death that vary by state, we also investigated whether this correlation was maintained while adding weeks to the time between admission and death (Figure S2). Additionally, as we show in Supplemental Figure 3, if plasma use is divided into quintiles from lowest using weeks to highest using weeks, a dose-response relationship between use of plasma and mortality is apparent.

**Figure 3.**
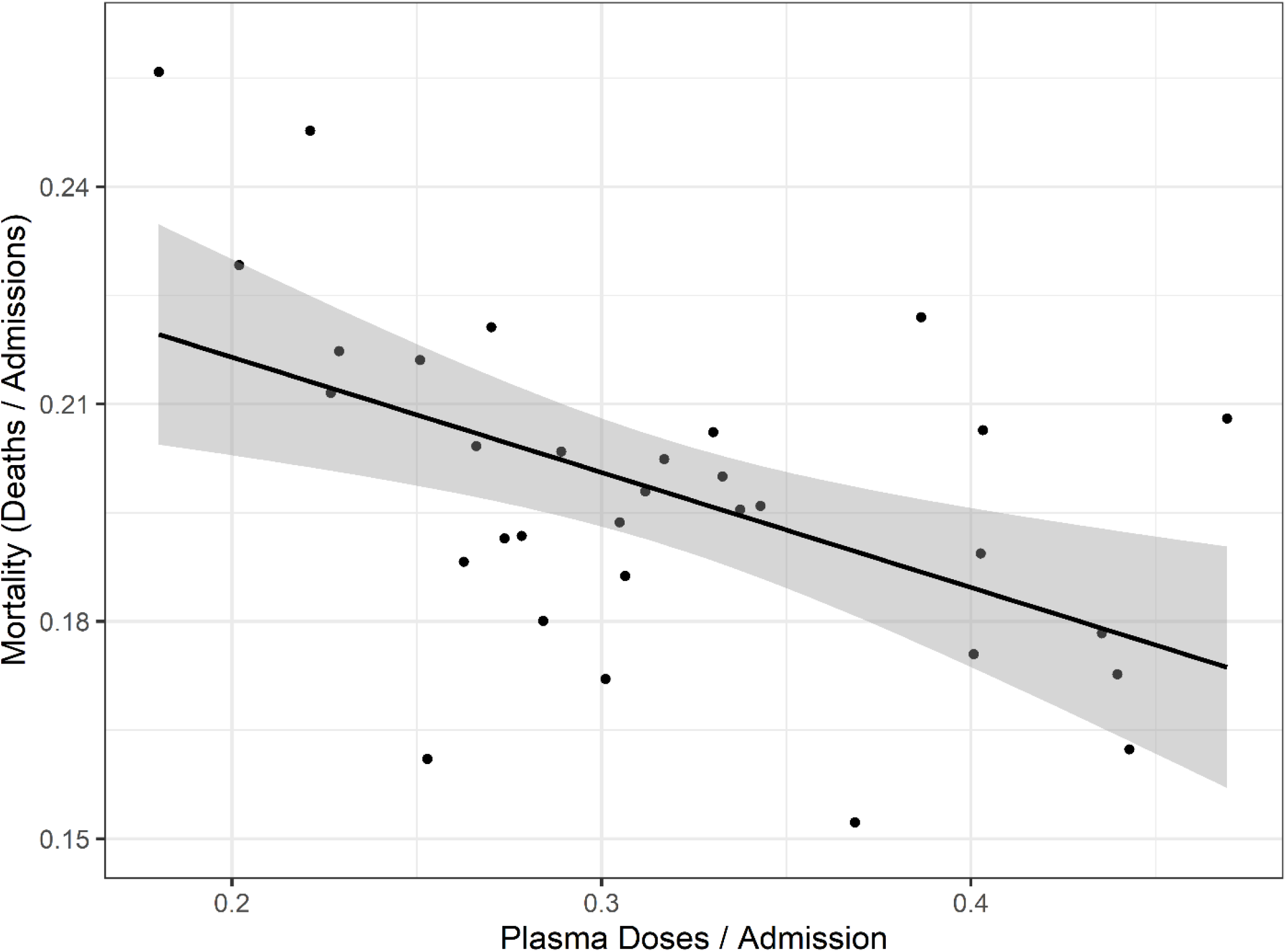
Correlation of mortality (death per admission) and CCP doses per admitted patients. Correlation analysis yields a Pearson’s correlation coefficient of −0.518 (p = 0.0024). The black line represents a linear model regression with an R squared of 0.268.

### Estimates of excess deaths

The linear model using the CCP utilization ratio to predict the CFR fit the data well in that this model explained 25% (R^2^ = 0.254) of the variance of CFR using only the CCP utilization ratio. The model estimated the CFR decreased by 1.8 percentage points for every 10-percentage point increase in the rate of utilization of CCP (p = 0.004). The linear regression analysis yielded a mortality in patients not receiving CCP of 25.2% as the y-intercept. A comparison of this number with that from USA studies shows reasonable agreement between with the average mortality of 23.5% in patients not receiving CCP (Table S3). This percentage also closely matches the 24% mortality for COVID-19 patients for the large RECOVERY trial in the United Kingdom ^12^ and the 30% mortality of patients receiving late CCP in analysis from the Expanded Access Program ^5^. An extrapolation of the linear model to the situation where every patient is treated with CCP yields a mortality of 7.6%, which is lower than the average mortality in USA studies, but still within the range reported (Table S3). However, this extrapolation to maximal use is much less reliable given the absence of points in the y axis region above an CCP utilization ratio of 0.6 and uncertainty as to whether the relationship is linear in those ranges. Hence, we caution the reader about making any strong inference from this estimate while noting that it is close to the 6.2% mortality reported for COVID-19 patients treated with high titer CCP very early upon hospitalization ^25^. Nevertheless, it is possible to use these efficacy numbers to estimate what the effect on deaths would have been had the United States continued to use CCP at the height of its usage in early Fall 2020, when more than 40% of all patients received plasma therapy.

With this model as a framework for estimating the excess number of deaths, the results of three scenarios were obtained (Figure 4). The total observed deaths over the study period was 356,534. Had the rate of CCP utilization observed during August through October 2002 carried over for the remaining months, the expected number of deaths was 327,516 (95%: CI: 293,811 to 361,221), which would result in 29,018 (95% CI: 3535 to 54,501) fewer deaths than observed. This excess death, in comparison, was small relative to the estimate that results from assuming plasma utilization was as high as it was after the EUA was issued. Had 50% utilization of plasma been continued, 62,383 (95% CI: 7599 to 117,166) fewer deaths may have been observed. Under the most extreme scenario comparing no plasma use to the highest observed use, a difference of 158,409 (95% CI: 19,296 to 297,523) deaths is estimated. We repeated this analysis independently for each of the databases used (CDC vs OWID) with concordant results (Figure S5 and Tables S1, S2).

**Figure 4.**
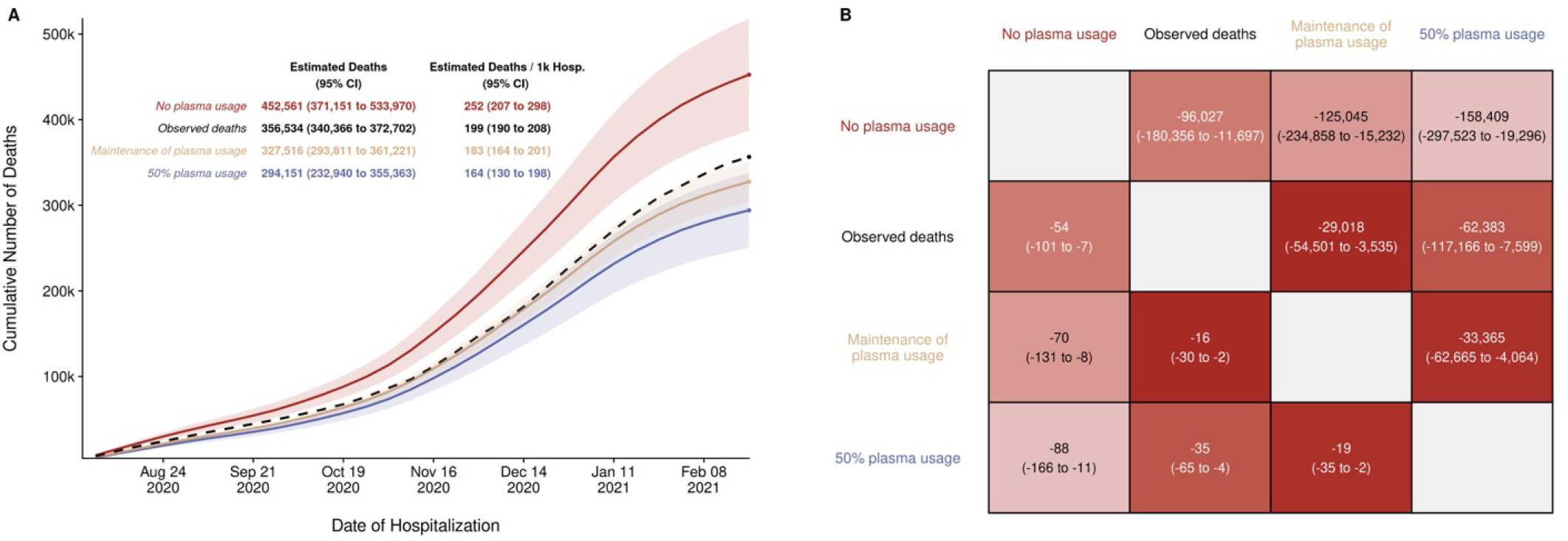
Estimated deaths under modeled scenarios of Covid-19 Convalescent Plasma. Panel A presents the longitudinal observed (dashed line) and modeled number of deaths under three scenarios for CCP over the study period (8/3/2020 – 2/22/2021) that included 356,534 deaths in 1,793,502 hospitalized patients. Over the entire study period, the CCP utilization ratio was 29.1%. In the scenario labeled maintenance of plasma, the CCP utilization ratio was set to 39.5%. With the no plasma and 50% plasma usage scenarios, the CCP utilization ratio was set at 0% and 50%, respectively. Panel B provides the pairwise comparisons of these scenarios to estimate the difference in expected number of deaths among the scenarios for the entire hospitalized patients (upper right triangle) and re-indexed to events per 1000 patients (lower left triangle). The rows represent the comparator or reference scenario, columns indicate the altered CCP use scenario. For example, the cell that intersects the observed deaths and the maintenance of plasma column shows that 29,018 fewer deaths would result had plasma use remained at the 39.5% level.

As an alternative method to estimating the excess deaths, which alleviated the need for a model, the data were summarized into two key periods (Table 1). In each of the seven weeks from September 21st to November 8th, 2020, the estimated proportion of in-patients transfused with convalescent plasma 42.6%. The two-week lagged mortality ratio for hospitalized patients was 18.16%. In the following period from 11/9/2021 to the week starting 3/22/21, transfusion rates declined steadily, averaging 27.4%. In parallel, mortality ratio rose to 20.08% for the 1,344,463 patients admitted in those 18 weeks. Table 1 shows that if the mortality rate in those 18 weeks had been the same as during the high-transfusion period, 25,871 fewer deaths would have taken place.

**Table 1.** Estimated number of excess deaths due to transfusion hesitancy.

| **Time Period** | **Transfusion Rate** | **Number of Admissions** | **Deaths** | **Mortality Rate** | **Expected deaths if mortality had remained at 18.16%** | **Excess deaths in the low transfusion period** | **Excess deaths per 1000 admissions** |
| --- | --- | --- | --- | --- | --- | --- | --- |
| High Utilization<br>(9/21/2020 - 11/8/2020) | 42.59% | 257,424 | 46,747 | 18.16% | 46,747 | -- | -- |
| Low Utilization<br>(11/9/2021 - 3/22/21) | 27.43% | 1,344,463 | 270,019 | 20.08% | 244,148 | 25,871 | 19.2 |

## Discussion

The use of CCP in hospitalized patients peaked from mid-September to the end of October 2020 shortly after the FDA allowed its widespread use under the EUA of August 23, 2020. This increase was not blunted by some claims that evidence of efficacy was insufficient for the authorization, ^26,27^ However, RCTs from India ^10^ and Argentina ^11^ reporting that CCP use did not reduce mortality in hospitalized patients were followed by a decline in use in November. This decline was accelerated after the RECOVERY trial in the UK issued a January, 2021 news release announcing no effect of CP on mortality ^12^. The pattern suggests that physicians were more influenced by clinical research data than by the controversies that played out in the media. A poll by the American Association of Blood Banks revealed a 50% increase in the number of institutions planning to stop offering CCP between February and March 2021, which cited lack of stronger efficacy data as the major reason for this decision ^28^.

The results show that CCP use per hospital admission closely paralleled the severity of the epidemic throughout much of 2020, increasing in parallel with hospital admissions, but then declined late in 2020 and in the early months of 2021, a time following the publication of several negative RCT studies. Of concern, the drop in per capita CCP utilization appeared to be associated with an increase in mortality among hospitalized COVID-19 patients. If there is a causal link between these two trends, we estimate that the decline in per capita CCP use resulted in 29,000-36,000 excess deaths. Given that this analysis was done during the ongoing pandemic and that the variables that affect mortality are not fully understood we urge caution with our findings. Nevertheless, applying Occam’s razor to the problem, the results do raise the disturbing possibility that reduced CCP use may have contributed to the deaths of thousands of patients.

While the relationship we describe needs to be interpreted with caution, several aspects of the evidentiary landscape point to CPP efficacy when used appropriately.

- An analysis of several dozen studies for which results were available by mid-January 2021 associated the administration of CCP with reduced mortality, with an effect size of ∼35% ^29^.
- Convalescent plasma has proven effective in individuals with defective humoral immunity and B cell defects ^30,31^.
- The active agent in plasma, antibody titer, is strongly related to mortality in transfused patients ^5,13,32^
- The active agent of CCP, specific antibody to SARS-CoV-2, has powerful antiviral activity and has been shown to reduce the viral load in patients with COVID-19, thus providing a mechanistic explanation for its therapeutic effect ^33^.
- Human convalescent plasma is protective in murine models of COVID-19 ^34^.
- Patients treated with CCP manifest reduced inflammatory markers ^35-37^ that can lead to host damage. ^38^
- Research has identified nine places where plasma affects the cascade of inflammation to the benefit of treated patients ^39^.

Late use of plasma, or use in people with advanced illness, is not likely to be effective, because at that time, the inflammatory response itself is the major pathophysiologic pathway to severe illness and death, by which time antibody is powerless to change the course of illness. ^40^

The results of RCTs are mixed, but many trials treated very advanced disease. For example, while the overall findings of the RECOVERY trial were null ^12^, patients treated early in the course of illness (before antibody conversion, in the first seven days, not receiving steroids or oxygen) all showed lower mortality in the CPP arm.

The observed association between CCP use per hospitalized patient and mortality helps define the potential for effectiveness of this therapy at a population level. This inference is strengthened by the fact that mortality from COVID-19 among hospitalized patients decreased substantially over most of 2020, consistent with worldwide trends ^19^, but then began to rise in late November and early December 2020, a period that coincided with precipitous reduction in CCP/admission.

Buttressing such a conclusion would require excluding the contribution of other variables that can affect mortality, which is challenging during an ongoing pandemic where information about the pathophysiology and clinical course of COVID-19 is accruing rapidly. Three factors can be excluded. The mean age of the hospital population with COVID-19 was actually older during peak use of CCP than later. The number of hospital admissions per week was not associated with increased mortality. The occasional delay seen in recording of death certificate information would apply largely to the most recent months when mortality was already highest. This analysis – based as it is on ecological correlations - must acknowledge some critical limitations. Many possible confounders are not available to evaluate for this analysis.

The emergence of SARS-CoV-2 variants with higher mortality is another variable that must be considered. In the United Kingdom a new lineage of SARS-CoV-2 emerged in the September 2020 known as B.1.1.7, which is associated with higher contagiousness and perhaps, mortality ^41^. This variant was identified in the United States in early winter 2021 but as of January only 76 cases had been described in 12 states, which was estimated to be <0.5% of the infections at the time ^42^. Hence this variant constituted a small minority of cases during much of the time involved in our analysis. Even as late as mid-March 2021 the CDC estimated that B.1.1.17 variant comprised only 25% of US isolates and increased mortality from these infections would not manifest itself until times later than our analysis ^43^. Limiting our analysis to the time from March 2020 to January 2021 shows maintenance of the trends described here.

Taking a page from the reluctance of citizens to accept vaccines against SARS-CoV-2, a phenomenon that has been termed ‘vaccine hesitancy’ ^44^, we call the phenomenon of reduced use of CCP ‘plasma hesitancy’. In this regard, we note that both the vaccines being administered in the USA and CCP are being used under a EUA regulatory framework, since neither has full approval status and that these hesitancies lead to avoidance behaviors based on the interpretation of available data by the public and health care providers, respectively. Plasma hesitancy may be a result of health care providers overvaluing and over-emphasizing negative results from RCT findings while dismissing other evidence that CCP reduces mortality. On the other hand, remdesevir has been widely embraced by the health care provider community for treatment of Sars-CoV2, despite a lack published data on reductions in mortality from its use and with conflicting RCT data regarding its clinical usefulness ^45,46^. In contrast, CCP has some data demonstrating a reduction in mortality for hospitalized patients with COVID-19 from its early use in non-ventilated patients^29^, and clear evidence of effectiveness in outpatients^13^ and yet has met with use hesitancy.

Why might this be? One possibility is that there is relatively little recent experience with treating infectious diseases with antibody therapies ^40^. The different receptions for plasma and for chemical agents could reflect confirmational bias by a health care community experienced in the successful use of antiviral drugs against Human immunodeficiency (HIV) and Hepatitis C (HCV) viruses.

In summary, CCP is one of few options available to physicians for treating COVID-19. It is available wherever there are recovered patients, it is relatively inexpensive and has a good safety profile ^3^. With regards to its efficacy, we have argued that physicians need to consider all the evidence, from observational studies to RCTs, when evaluating clinical efficacy data ^47^. We note that the FDA reaffirmed the EUA status of CCP in February 2021 ^2^ by permitting its continued use in hospitalized patients if used early in COVID-19 and with units that have a sufficient content of specific antibody. In addition, interim guidelines for American Association of Blood Banks ^48^ and Brazil ^49^ emphasize that CCP is more likely to be effective when used in early COVID-19 with units having high content of specific antibody. We are hopeful that physicians consider the totality of the available evidence, including our findings, when making decisions for CCP use in individual patients.

## Data Availability

All our data is save into files and is available.

## Acknowledgements

We are grateful to Bill Block and Jennifer Kapral from Blood Centers of America, Inc. (West Warwick, RI) and Kate Fry from America Blood Centers (Washington, DC) for their help in obtaining the plasma usage data. AC was supported in part by RO1 HL059842 and R01 AI152078 9; MJJ was supported in part by 5R35HL139854. This project has been funded in whole or in part with Federal funds from the Department of Health and Human Services; Office of the Assistant Secretary for Preparedness and Response; Biomedical Advanced Research and Development Authority under Contract No. 75A50120C00096.

## Disclaimer

*The views and opinions expressed in this publication are those of the authors and do not reflect the official policy or position of the US Department of Health and Human services and its agencies including the Biomedical Research and Development Authority and the Food and Drug Administration, as well as any agency of the U*.*S. government. Assumptions made within and interpretations from the analysis are not reflective of the position of any U*.*S. government entity*.

---

**Table S1.** Data from and calculations for excess mortality from CCP hesitancy based on the OWID database.

| **Week** | **Plasma Doses** | **Deaths** | **Hospitalized Patients** | **Admissions** | **Plasma Doses Per Patient** | **Mortality (Deaths / Admissions - 2 Week Shift)** |
| --- | --- | --- | --- | --- | --- | --- |
| 7/18/2020 | 11231 | 5559 | 38045 | 21481 | 0.52 | 0.37 |
| 7/25/2020 | 12919 | 6513 | 42802 | 33434 | 0.39 | 0.22 |
| 8/1/2020 | 11724 | 7855 | 49972 | 38256 | 0.31 | 0.19 |
| 8/8/2020 | 10511 | 7421 | 41313 | 33153 | 0.32 | 0.20 |
| 8/15/2020 | 10495 | 7126 | 38655 | 31543 | 0.33 | 0.20 |
| 8/22/2020 | 9218 | 6711 | 34466 | 30226 | 0.30 | 0.19 |
| 8/29/2020 | 8968 | 6309 | 31031 | 29776 | 0.30 | 0.17 |
| 9/5/2020 | 8424 | 5855 | 28242 | 27010 | 0.31 | 0.20 |
| 9/12/2020 | 7894 | 5124 | 26237 | 28351 | 0.28 | 0.19 |
| 9/19/2020 | 9647 | 5348 | 25382 | 23924 | 0.40 | 0.21 |
| 9/26/2020 | 11670 | 5439 | 25915 | 24867 | 0.47 | 0.21 |
| 10/3/2020 | 11101 | 4939 | 27804 | 27575 | 0.40 | 0.19 |
| 10/10/2020 | 14796 | 5172 | 31370 | 33658 | 0.44 | 0.17 |
| 10/17/2020 | 16401 | 5222 | 34934 | 37027 | 0.44 | 0.16 |
| 10/24/2020 | 17827 | 5812 | 39899 | 40935 | 0.44 | 0.18 |
| 10/31/2020 | 18628 | 6012 | 45983 | 46467 | 0.40 | 0.18 |
| 11/7/2020 | 19205 | 7303 | 55640 | 56877 | 0.34 | 0.20 |
| 11/14/2020 | 26176 | 8156 | 69432 | 70999 | 0.37 | 0.15 |
| 11/21/2020 | 28076 | 11119 | 81702 | 81834 | 0.34 | 0.20 |
| 11/28/2020 | 28688 | 10814 | 92351 | 86841 | 0.33 | 0.21 |
| 12/5/2020 | 27350 | 16033 | 98549 | 94573 | 0.29 | 0.20 |
| 12/12/2020 | 25706 | 17907 | 107227 | 101624 | 0.25 | 0.16 |
| 12/19/2020 | 29842 | 19238 | 112272 | 104959 | 0.28 | 0.22 |
| 12/26/2020 | 24350 | 16363 | 118892 | 106311 | 0.23 | 0.22 |
| 1/9/2021 | 30576 | 23074 | 126420 | 116363 | 0.26 | 0.19 |
| 1/16/2021 | 28946 | 23726 | 120658 | 108721 | 0.27 | 0.20 |
| 1/23/2021 | 25395 | 21843 | 106881 | 93972 | 0.27 | 0.22 |
| 1/30/2021 | 20226 | 22152 | 91286 | 80589 | 0.25 | 0.22 |
| 2/6/2021 | 18913 | 20667 | 77709 | 69080 | 0.27 | 0.19 |
| 2/13/2021 | 12303 | 17346 | 63237 | 55607 | 0.22 | 0.25 |
| 2/20/2021 | 8369 | 13155 | 53512 | 46421 | 0.18 | 0.25 |
| 2/27/2021 | 8429 | 13706 | 44799 | 41727 | 0.20 | NA |
| 3/6/2021 | 6006 | 11791 | NA | 32075 | 0.19 | NA |

**Table S2.** Replicate excess death calculations based on OWID database.

|  | Scenario 1 |  | Scenario 2 |  | Scenario 3 |  |
| --- | --- | --- | --- | --- | --- | --- |
| Transfused | 38.90% |  | 50% |  | 0% |  |
| Week | Expected Deaths | Difference | Expected Deaths | Difference | Expected Deaths | Difference |
| 8/1/2020 | 3974.94 | 3880.06 | 3625.35 | 4229.65 | 5332.66 | 1707.31 |
| 8/8/2020 | 6186.78 | 1234.22 | 5642.66 | 1778.34 | 8299.99 | 2657.33 |
| 8/15/2020 | 7079.06 | 46.94 | 6456.47 | 669.53 | 9497.05 | 3040.59 |
| 8/22/2020 | 6134.78 | 576.22 | 5595.23 | 1115.77 | 8230.23 | 2635.00 |
| 8/29/2020 | 5836.86 | 472.14 | 5323.51 | 985.49 | 7830.55 | 2507.04 |
| 9/5/2020 | 5593.16 | 261.84 | 5101.24 | 753.76 | 7503.60 | 2402.36 |
| 9/12/2020 | 5509.89 | -385.89 | 5025.30 | 98.70 | 7391.89 | 2366.60 |
| 9/19/2020 | 4998.05 | 349.95 | 4558.48 | 789.52 | 6705.23 | 2146.75 |
| 9/26/2020 | 5246.20 | 192.80 | 4784.80 | 654.20 | 7038.14 | 2253.34 |
| 10/3/2020 | 4427.00 | 512.00 | 4037.65 | 901.35 | 5939.13 | 1901.48 |
| 10/10/2020 | 4601.50 | 570.50 | 4196.80 | 975.20 | 6173.23 | 1976.43 |
| 10/17/2020 | 5102.60 | 119.40 | 4653.83 | 568.17 | 6845.49 | 2191.66 |
| 10/24/2020 | 6228.23 | -416.23 | 5680.46 | 131.54 | 8355.60 | 2675.14 |
| 10/31/2020 | 6851.64 | -839.64 | 6249.05 | -237.05 | 9191.95 | 2942.91 |
| 11/7/2020 | 7574.80 | -271.80 | 6908.60 | 394.40 | 10162.11 | 3253.51 |
| 11/14/2020 | 8598.46 | -442.46 | 7842.24 | 313.76 | 11535.43 | 3693.20 |
| 11/21/2020 | 10524.78 | 594.22 | 9599.13 | 1519.87 | 14119.72 | 4520.58 |
| 11/28/2020 | 13137.97 | -2323.97 | 11982.50 | -1168.50 | 17625.50 | 5643.00 |
| 12/5/2020 | 15142.93 | 890.07 | 13811.12 | 2221.88 | 20315.29 | 6504.17 |
| 12/12/2020 | 16069.45 | 1837.55 | 14656.16 | 3250.84 | 21558.28 | 6902.12 |
| 12/19/2020 | 17500.21 | 1737.79 | 15961.09 | 3276.91 | 23477.75 | 7516.66 |
| 12/26/2020 | 18804.96 | -2441.96 | 17151.08 | -788.08 | 25228.16 | 8077.08 |
| 1/9/2021 | 19422.09 | 3651.91 | 17713.93 | 5360.07 | 26056.07 | 8342.14 |
| 1/16/2021 | 19672.27 | 4053.73 | 17942.11 | 5783.89 | 26391.71 | 8449.60 |
| 1/23/2021 | 21532.33 | 310.67 | 19638.58 | 2204.42 | 28887.11 | 9248.53 |
| 1/30/2021 | 20118.22 | 2033.78 | 18348.84 | 3803.16 | 26989.99 | 8641.15 |
| 2/6/2021 | 17389.00 | 3278.00 | 15859.65 | 4807.35 | 23328.55 | 7468.89 |
| 2/13/2021 | 14912.55 | 2433.45 | 13601.01 | 3744.99 | 20006.22 | 6405.21 |
| 2/20/2021 | 12782.87 | 372.13 | 11658.63 | 1496.37 | 17149.11 | 5490.48 |
| 2/27/2021 | 10289.77 | 3416.23 | 9384.79 | 4321.21 | 13804.44 | 4419.64 |
| 3/6/2021 | 8589.95 | 3201.05 | 7834.47 | 3956.53 | 11524.01 | 3689.54 |
| **Excess Deaths** |  | **28904.69** |  | **57913.24** |  | **141669.44** |

**Table S3.**
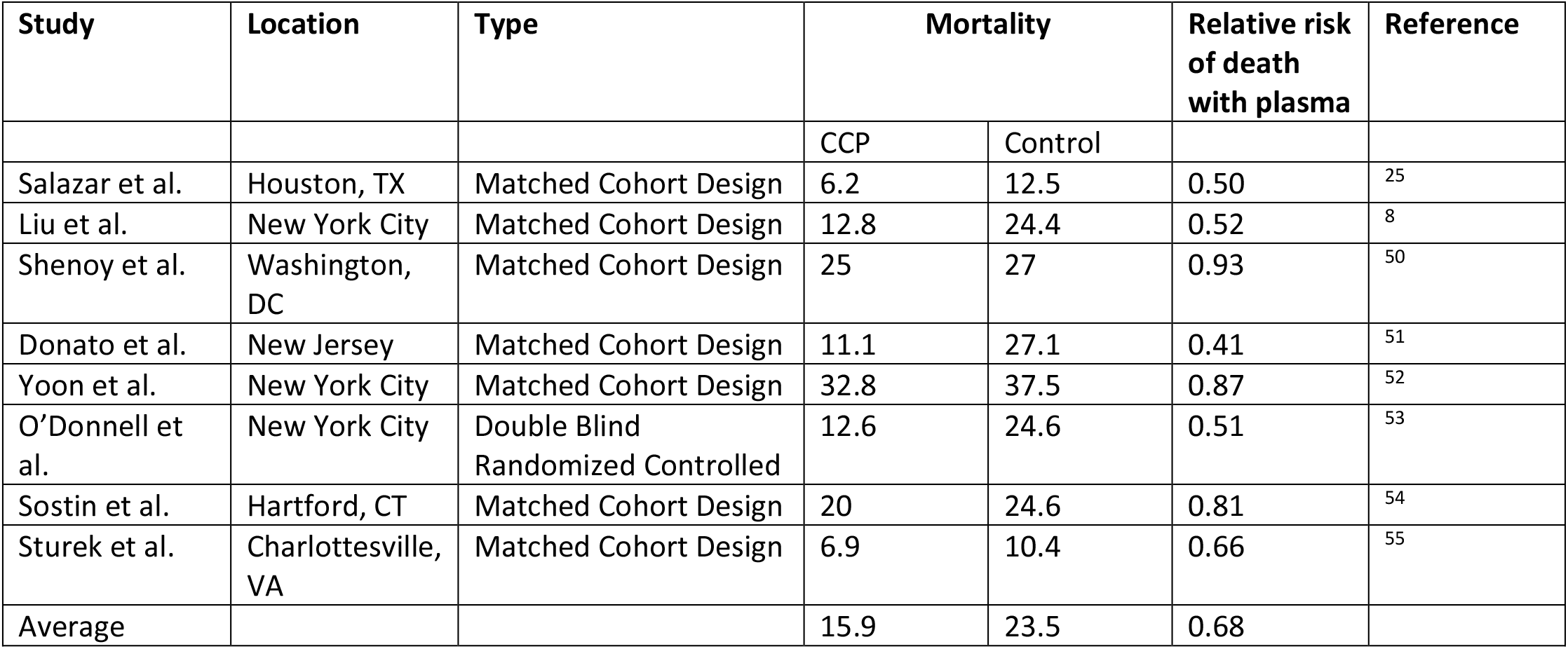
Mortality of COVID-19 in USA CCP efficacy studies.

**Table S4.**
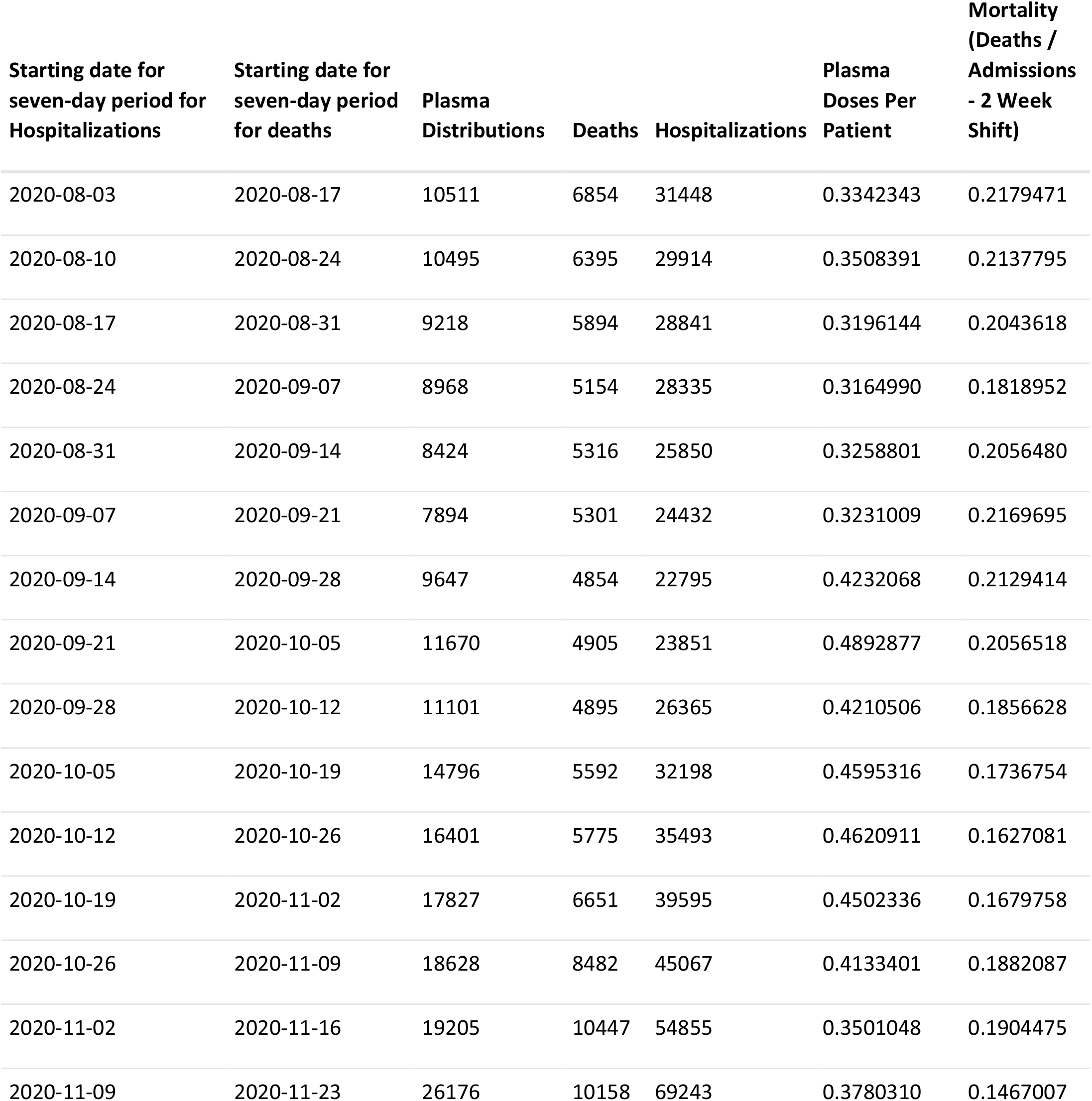
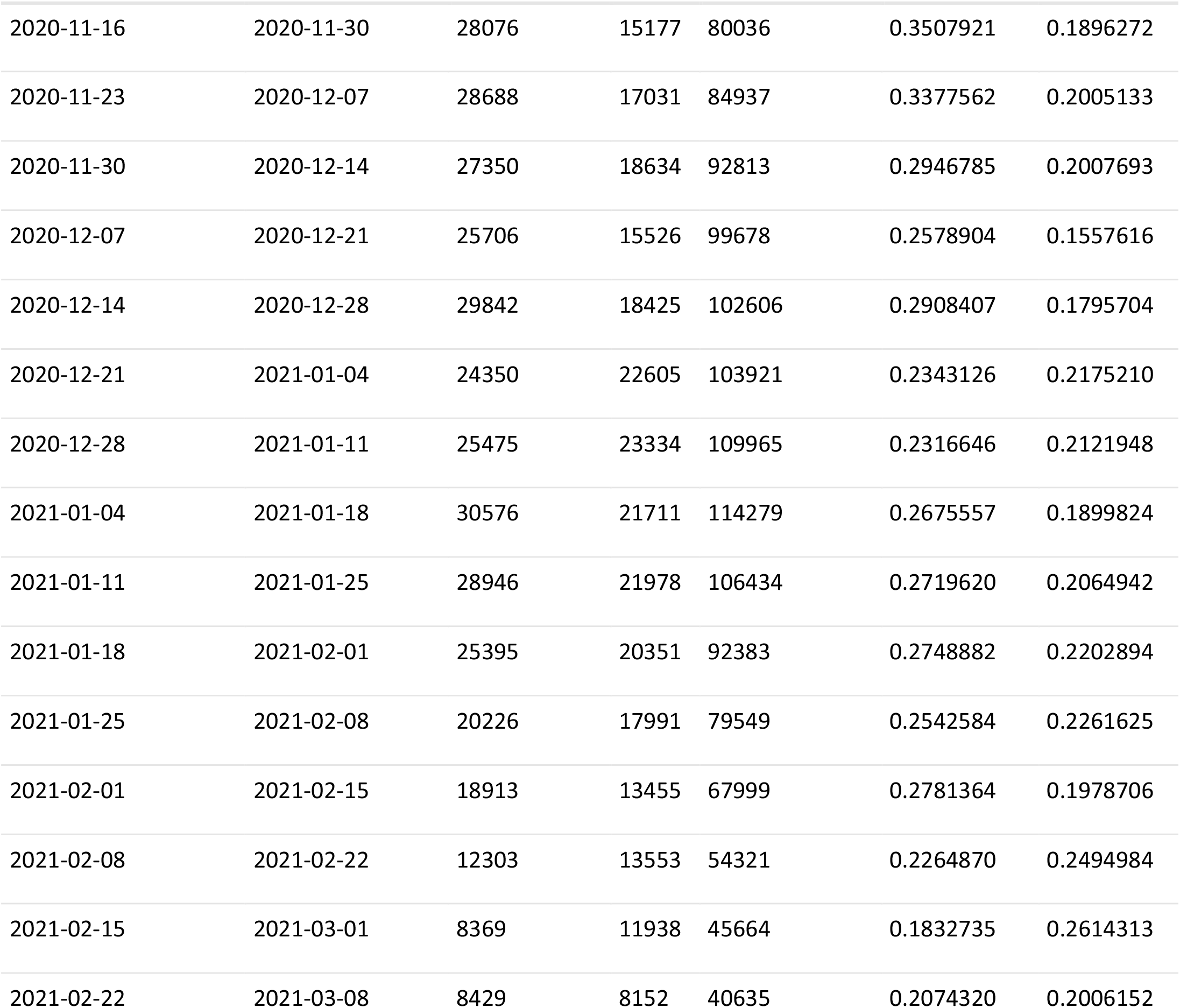
Data from and calculations for excess mortality from CCP hesitancy based on the CDC database.

| Starting date for seven-day period for Hospitalizations | Starting date for seven-day period for deaths | Plasma Distributions | Deaths | Hospitalizations | Plasma Doses Per Patient | Mortality (Deaths / Admissions - 2 Week Shift) |
| --- | --- | --- | --- | --- | --- | --- |
| 2020-08-03 | 2020-08-17 | 10511 | 6854 | 31448 | 0.3342343 | 0.2179471 |
| 2020-08-10 | 2020-08-24 | 10495 | 6395 | 29914 | 0.3508391 | 0.2137795 |
| 2020-08-17 | 2020-08-31 | 9218 | 5894 | 28841 | 0.3196144 | 0.2043618 |
| 2020-08-24 | 2020-09-07 | 8968 | 5154 | 28335 | 0.3164990 | 0.1818952 |
| 2020-08-31 | 2020-09-14 | 8424 | 5316 | 25850 | 0.3258801 | 0.2056480 |
| 2020-09-07 | 2020-09-21 | 7894 | 5301 | 24432 | 0.3231009 | 0.2169695 |
| 2020-09-14 | 2020-09-28 | 9647 | 4854 | 22795 | 0.4232068 | 0.2129414 |
| 2020-09-21 | 2020-10-05 | 11670 | 4905 | 23851 | 0.4892877 | 0.2056518 |
| 2020-09-28 | 2020-10-12 | 11101 | 4895 | 26365 | 0.4210506 | 0.1856628 |
| 2020-10-05 | 2020-10-19 | 14796 | 5592 | 32198 | 0.4595316 | 0.1736754 |
| 2020-10-12 | 2020-10-26 | 16401 | 5775 | 35493 | 0.4620911 | 0.1627081 |
| 2020-10-19 | 2020-11-02 | 17827 | 6651 | 39595 | 0.4502336 | 0.1679758 |
| 2020-10-26 | 2020-11-09 | 18628 | 8482 | 45067 | 0.4133401 | 0.1882087 |
| 2020-11-02 | 2020-11-16 | 19205 | 10447 | 54855 | 0.3501048 | 0.1904475 |
| 2020-11-09 | 2020-11-23 | 26176 | 10158 | 69243 | 0.3780310 | 0.1467007 |

| Starting date for<br>seven-day period for<br>Hospitalizations | Starting date for<br>seven-day period<br>for deaths | Plasma<br>Distributions | Deaths | Hospitalizations | Plasma<br>Doses Per<br>Patient | Mortality<br>(Deaths /<br>Admissions<br>- 2 Week<br>Shift) |
| --- | --- | --- | --- | --- | --- | --- |
| 2020-11-16 | 2020-11-30 | 28076 | 15177 | 80036 | 0.3507921 | 0.1896272 |
| 2020-11-23 | 2020-12-07 | 28688 | 17031 | 84937 | 0.3377562 | 0.2005133 |
| 2020-11-30 | 2020-12-14 | 27350 | 18634 | 92813 | 0.2946785 | 0.2007693 |
| 2020-12-07 | 2020-12-21 | 25706 | 15526 | 99678 | 0.2578904 | 0.1557616 |
| 2020-12-14 | 2020-12-28 | 29842 | 18425 | 102606 | 0.2908407 | 0.1795704 |
| 2020-12-21 | 2021-01-04 | 24350 | 22605 | 103921 | 0.2343126 | 0.2175210 |
| 2020-12-28 | 2021-01-11 | 25475 | 23334 | 109965 | 0.2316646 | 0.2121948 |
| 2021-01-04 | 2021-01-18 | 30576 | 21711 | 114279 | 0.2675557 | 0.1899824 |
| 2021-01-11 | 2021-01-25 | 28946 | 21978 | 106434 | 0.2719620 | 0.2064942 |
| 2021-01-18 | 2021-02-01 | 25395 | 20351 | 92383 | 0.2748882 | 0.2202894 |
| 2021-01-25 | 2021-02-08 | 20226 | 17991 | 79549 | 0.2542584 | 0.2261625 |
| 2021-02-01 | 2021-02-15 | 18913 | 13455 | 67999 | 0.2781364 | 0.1978706 |
| 2021-02-08 | 2021-02-22 | 12303 | 13553 | 54321 | 0.2264870 | 0.2494984 |
| 2021-02-15 | 2021-03-01 | 8369 | 11938 | 45664 | 0.1832735 | 0.2614313 |
| 2021-02-22 | 2021-03-08 | 8429 | 8152 | 40635 | 0.2074320 | 0.2006152 |

**Supplementary Figure 1.**
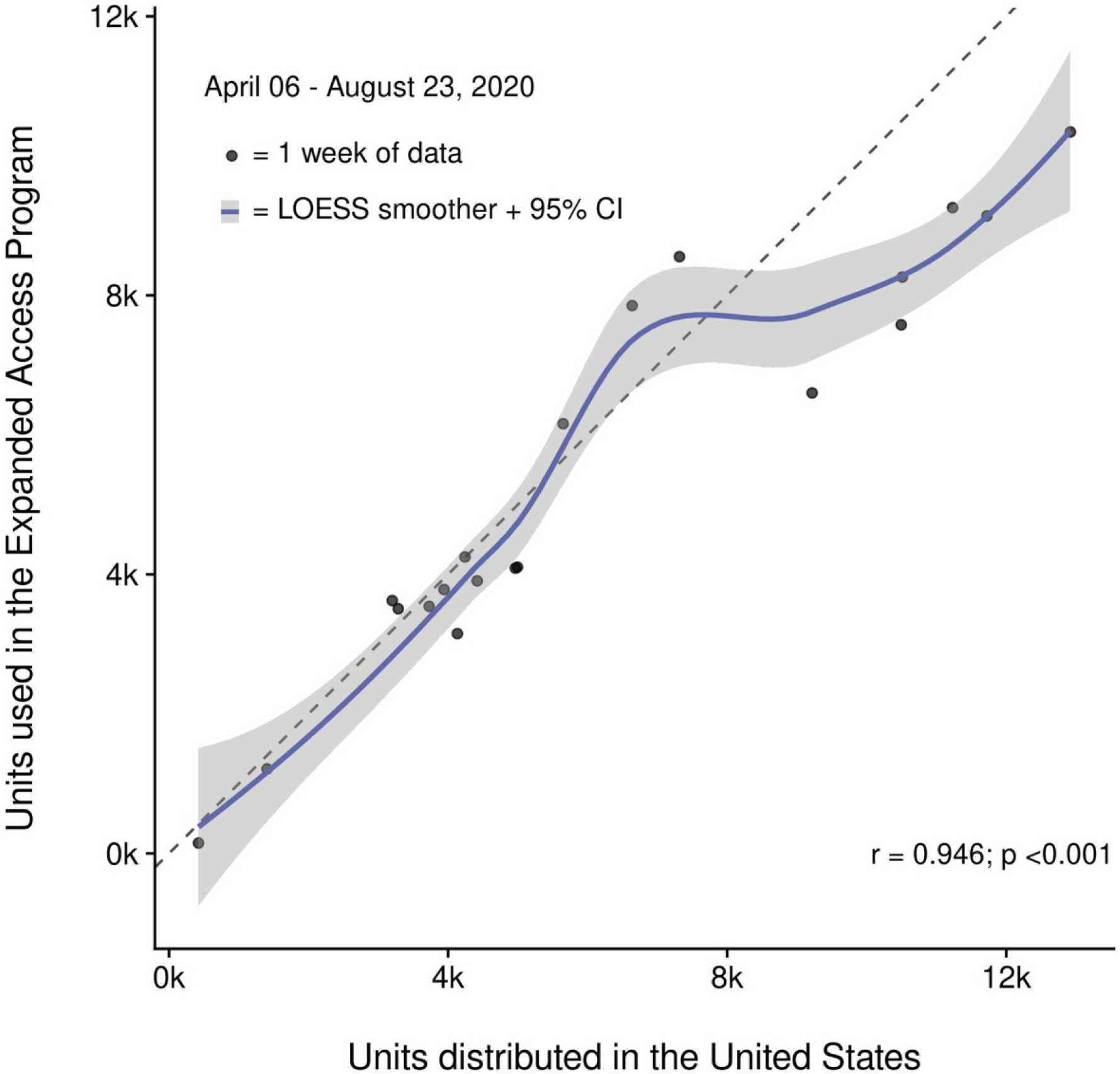
Correlation of convalescent plasma distribution and usage within the Expanded Access Program. Shown is the progressive increase in the number of convalescent plasma units distributed in the United States and convalescent plasma units used in the Expanded Access Program (EAP). Data between April 06 and August 23, 2020 are pooled in weekly intervals and represented as filled circles. The Pearson correlation coefficient was used to assess correlation (r= 0.946, P<0.001) and a LOESS smoother with a 95% confidence interval (CI) and a reference line were overlayed. Points below the reference line represent weeks where more convalescent plasma was distributed than used within the EAP. Conversely, points above the reference line are indicative of more convalescent plasma being used in the EAP than distributed.

**Supplemental Figure 2.**
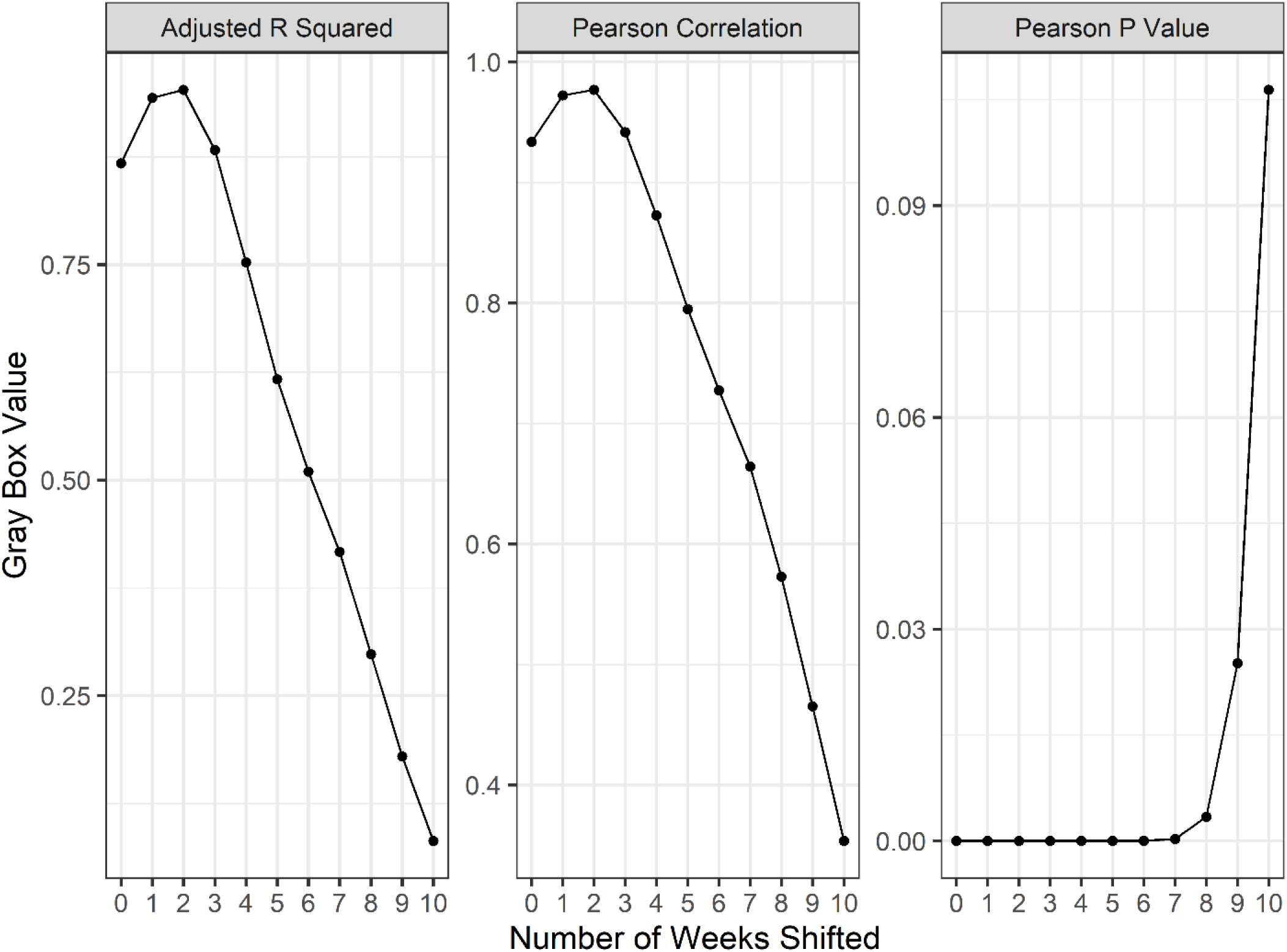
A series of linear regressions and Pearson’s correlation tests comparing weekly reported deaths to new weekly hospital admissions, offset by various numbers of weeks to identify the length of lag between admission and death of patients. Y axes values reflect the parameter of each gray box throughout the shifted weeks. Correlations peak at 2-3 weeks shifted, suggesting the lag time between admission and reported death is roughly two weeks

**Supplemental Figure 3.**
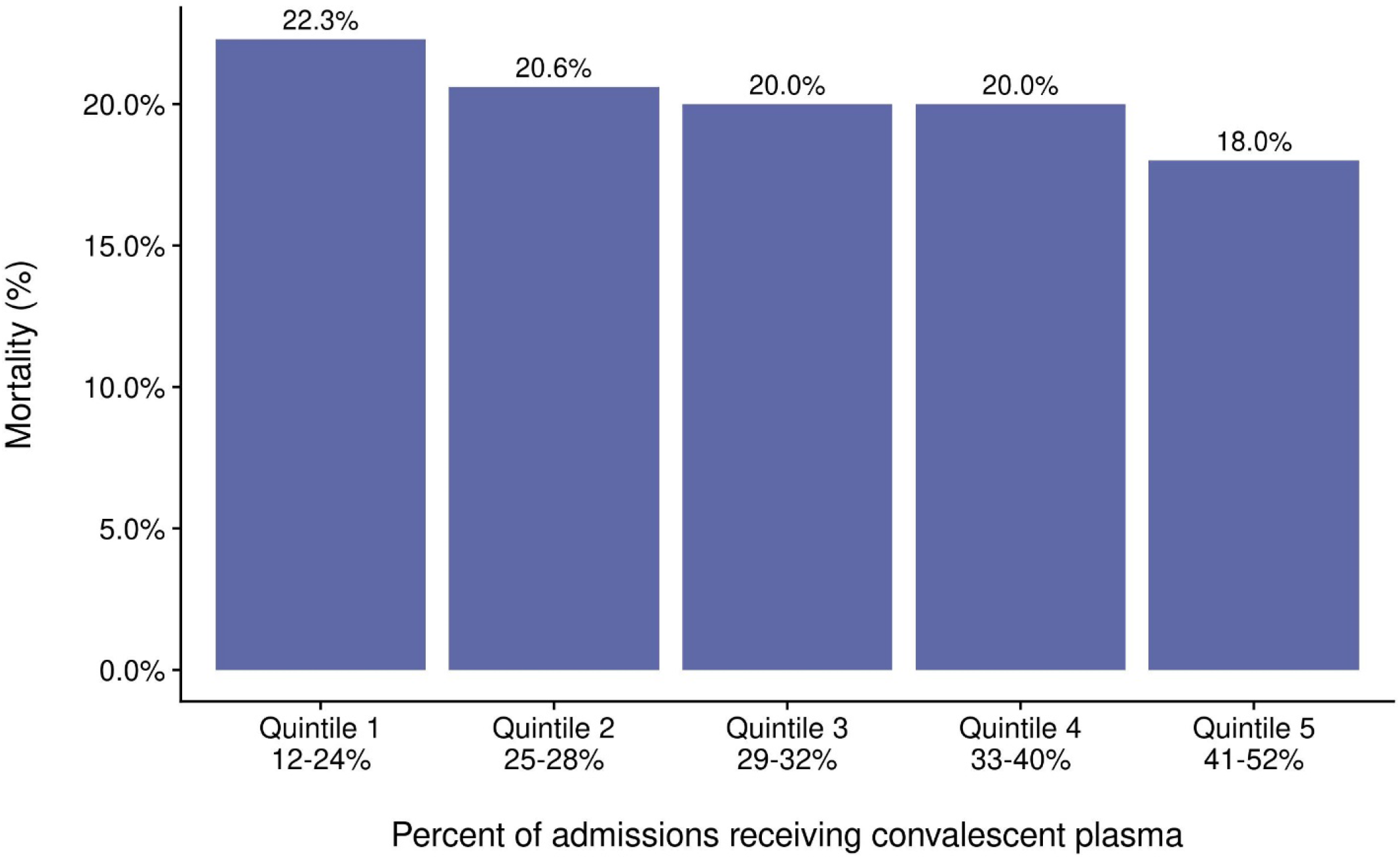
Mortality from COVID-19 by quintile of percent of admissions receiving CCP.

**Supplemental Figure 4.**
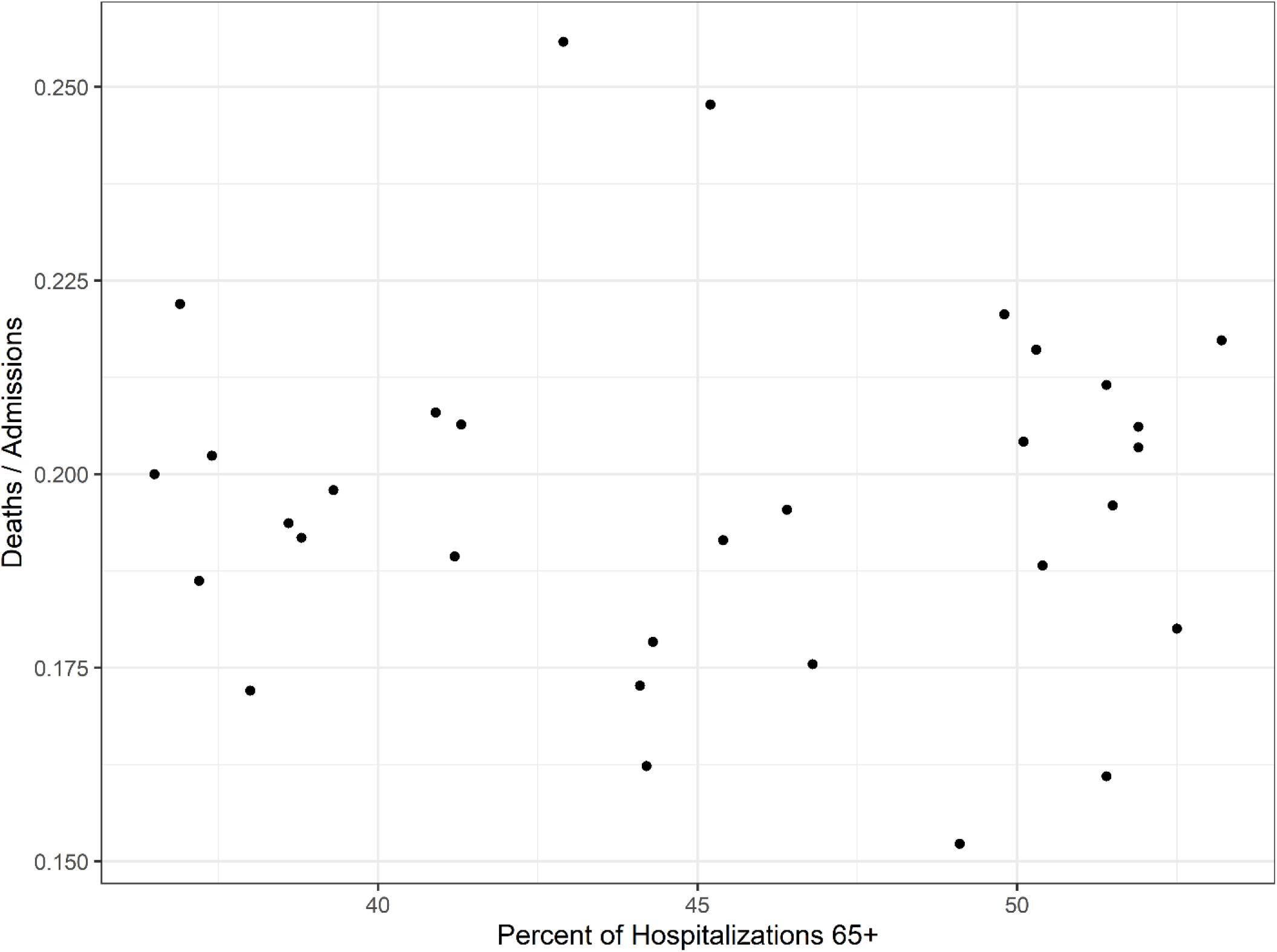
Investigation of high age group mortality. The shifted mortality is compared to the percent of hospitalized patients 65+ each week as reported by the CDC. There is no significant correlation between the two variables, suggesting changes in mortality are not explainable by an increase in hospitalized high risk patients.

**Supplemental Figure 5.**
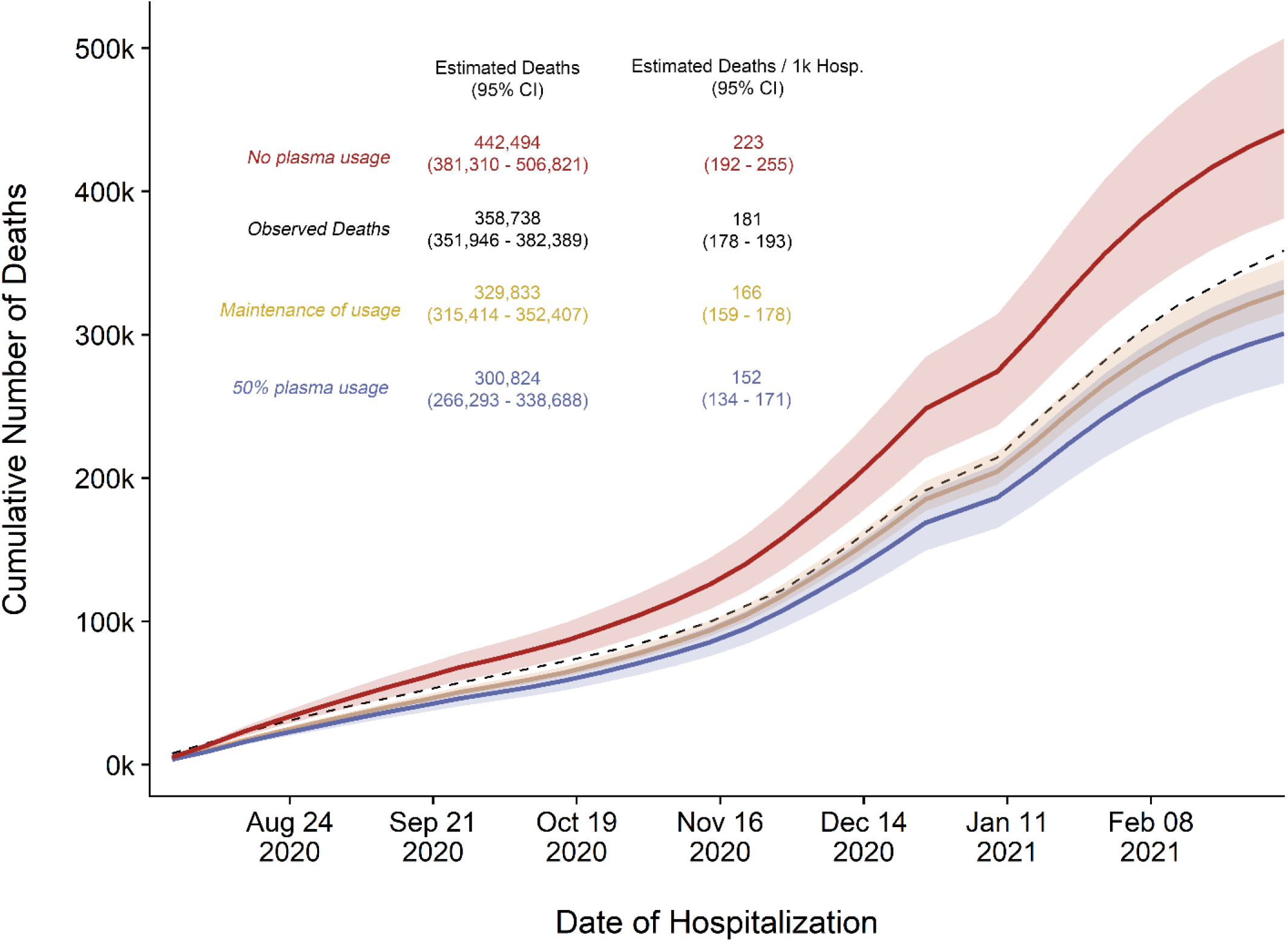
Replicated cumulative excess deaths analysis per OWID database for scenario 1 (orange): Maintained plasma transfusion rate from Oct-Nov throughout period, scenario 2 (blue): 50% transfusion rate throughout period, and scenario 3 (red): 0% transfusion rate throughout period. Black dashed line represents observed cumulative deaths per OWID reporting.

